# Vestibular patients exhibit drawing and writing size distortions similar to astronauts during spaceflight

**DOI:** 10.1101/2025.07.26.25332244

**Authors:** Gilles Clément, Olivier Deguine

## Abstract

This study aimed to examine how vestibular dysfunction influences spatial perception and motor output, using drawing tasks as a tool to explore changes in body schema. Patients with otolith dysfunction were compared to astronauts tested before, during, and after six months of exposure to microgravity aboard the International Space Station. Participants were instructed to draw and write words both horizontally and vertically. When performing these tasks with eyes closed, vestibular patients frequently produced drawings and handwriting that were more vertically compressed compared to when their eyes were open. Similarly, astronauts exhibited greater vertical compression in drawings and handwriting during spaceflight compared to their preflight performance. These findings highlight the critical role of vestibular input in integrating multisensory information to preserve consistent spatial representations.

## Introduction

To maintain balance and spatial orientation, the brain must integrate multiple sources of sensory information— vestibular, visual, and proprioceptive signals. This integration creates an internal map or “body schema” and a mental representation of the surrounding environment. The sensorimotor system contributes not only to balance and spatial orientation but also to the accurate perception of scale and body schema—how large or small objects (including one’s own body parts) are represented mentally [1].

Because drawing engages complex sensorimotor coordination and cognitive processing, analyzing how someone draws can reveal how well their brain integrates sensory inputs. The vestibular system plays a crucial role in detecting head movement and orientation relative to gravity. When vestibular function is compromised—due to injury, disease, or altered gravitational environments—this spatial orientation can become distorted. These distortions often reveal themselves in subtle ways: for example, a patient might consistently draw figures leaning to one side, asymmetrical shapes, or write words that might slant abnormally [2,3]. Studies using line-drawing tasks with vestibular stimulation have shown that head tilt or galvanic vestibular stimulation can led to rotated or skewed drawings, reflecting an underlying imbalance in spatial perception and hand movement control [4].

This study aimed to investigate how vestibular dysfunction–whether in patients on Earth or astronauts in microgravity–affects spatial perception using drawing and writing tasks to probe alterations in sensorimotor integration. One group of 20 vestibular patients (15 women, 5 men; aged 34-65 years) was selected according to the following signs and symptoms: (a) absence of rotatory nystagmus; (b) abnormal vestibular-evoked myographic potentials; (c) postural instability; (d) actual or subjective deviation from the straight path while walking; (e) perceived tilt or distortion of the body or the environment; (f) erroneous sensation of linear motion during transportation on foot, by car, train, aircraft, elevator, etc.; and (g) floating or rising sensation. These symptoms are indicative of a vestibular disorder involving otolith dysfunction [5].

## Material and Methods

Another group of 8 astronauts (one woman, 7men; aged 45-56 years) was tested before, during, and after long-duration missions on board the ISS. Mission durations ranged from 57 to 195 days (mean 154 days). Astronauts were all tested 3 times pre-flight (at approximately 90, 60, and 30 days before launch), 4 times in-flight (at approximately every month) and 3 times post-flight (just after lending, then 4 days and 8 days later).

All procedures were performed in compliance with relevant laws and institutional guidelines and have been approved by the *Comité de Protection des Personnes Sud-Ouest et Outre Mer* and NASA Institutional Review Board Committee for the Protection of Human Subjects. Privacy rights of human subjects have been observed and written informed consent was obtained for experimentation with astronauts and vestibular patients.

Vestibular patients were asked to draw squares, crosses, and circles, and to write the word “marguerite” horizontally and vertically on paper, first with the eyes open and then with the eyes closed (Fig. 1). Astronauts performed a similar task while wearing a head-mounted display (Z800 3DVisor, eMagin Corporation, Bellevue, WA, USA) while wearing a shroud to eliminate all external visual cues. Using an electronic pen and a digital tablet (Intuos A4; Wacom Co., Ltd, Vancouver, WA, USA; active area: 305 × 231 mm, resolution: 5080 lpi), they drew the same shapes and wrote the word “microgravity” without visual feedback. Preliminary results obtained on squares and crosses drawn by astronauts have been reported in a previous publication [6].

**Fig. 1.**
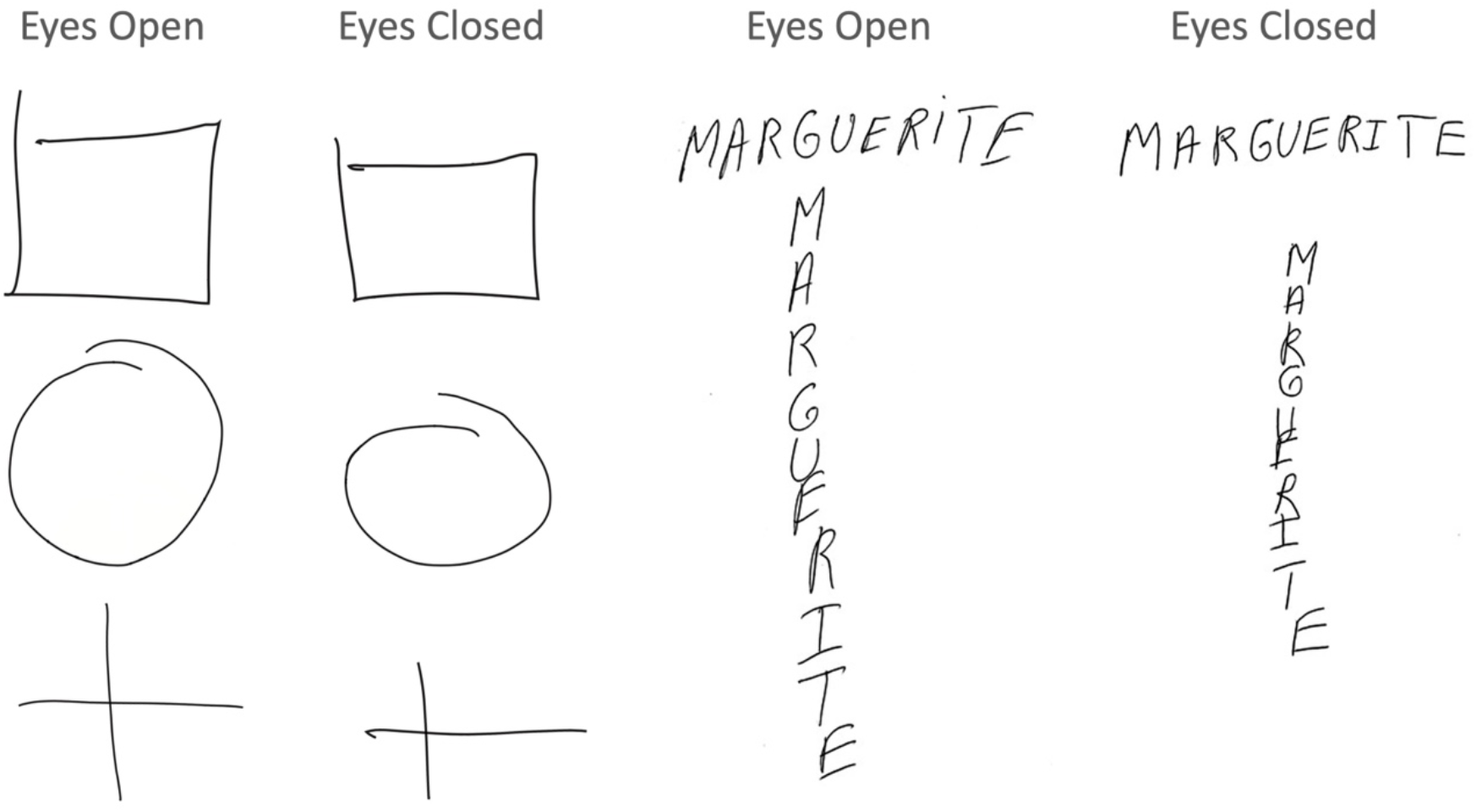
Examples of drawings and writings by one vestibular patient with eyes open and eyes closed.

Participants were instructed to match their drawings to their internal representation of a perfect square, circle, or cross. On Earth, vestibular patients and astronauts completed the task while seated. In-flight, the astronauts were free-floating, with the tablet secured to their thighs using knee straps. Each shape or word was drawn or written 6 times, with no time limit imposed.

For each trial, the horizontal and vertical dimensions of all shapes and written words were measured individually. The mean vertical-to-horizontal length ratio was then calculated for each shape, word, and condition (eyes open vs. eyes closed for vestibular patients; preflight, inflight, and postflight phases for astronauts). Astronaut data were grouped by flight phase. Paired (dependent) two-tailed t-tests were used to compare measurements within individuals across different visual conditions (vestibular patients) or flight phase (astronauts), with statistical significance set at p < 0.05.

## Results

In vestibular patients, the vertical-to-horizontal length ratio significantly decreased when drawings and writings were performed with eyes closed compared to eyes open. This reduction was consistent across all shapes: squares (t(19) = –4.09, p < 0.001), circles (t(19) = –4.15, p < 0.001), crosses (t(19) = –3.22, p < 0.01), and words (t(19) = –3.62, p < 0.01) (Fig. 2A).

**Fig. 2.**
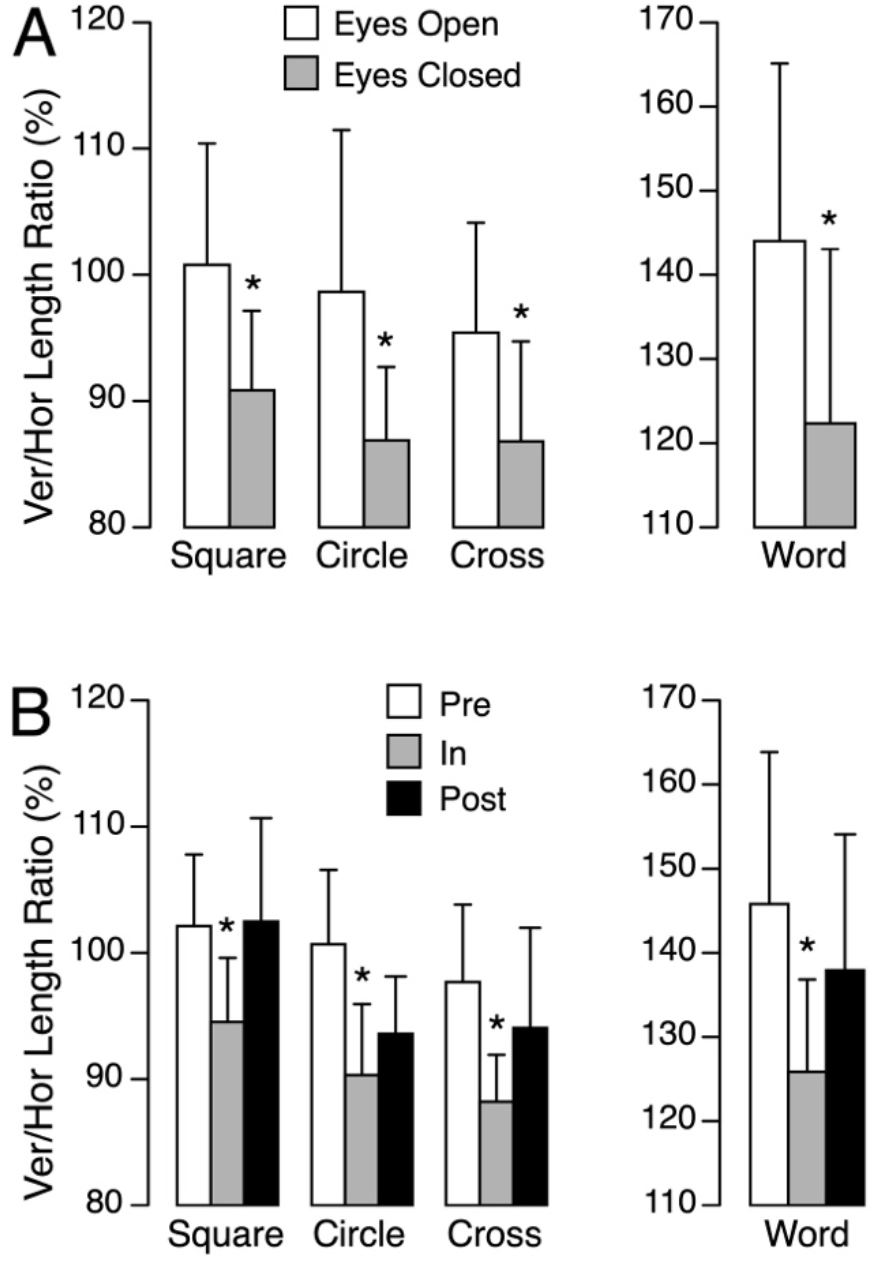
**A**. Comparison of the mean and standard deviation of the vertical-to-horizontal length ratio for drawings (squares, circles, and crosses) and for handwriting by vestibular patients, performed with eyes open and eyes closed. **B**. Comparison of the same ratio in astronauts performing the tasks with eyes closed before (Pre), during (In), and after (Post) a 6-month spaceflight. * p < 0.05.

A similar pattern emerged in astronauts. During spaceflight, the vertical-to-horizontal ratio significantly decreased compared to preflight measurements for all tasks: square (t(7) = –2.63, p = 0.034), circle (t(7) = –3.46, p = 0.011), and cross (t(7) = –3.05, p = 0.019) writing, as well as word writing (t(7) = –6.23, p < 0.001). Across all shapes, the vertical-to-horizontal length ratio was, on average, 11.8 ± 5.8% lower during the flight compared to before the flight (Fig. 2B). After flight, the vertical-to-horizontal ratio returned to pre-flight levels. The vertical compression of spatial output while in microgravity mirrors the distortions seen in vestibular patients tested without vision. The consistency of this effect across both populations— vestibular-impaired patients on Earth and healthy individuals in the altered sensory environment of space—strongly suggests that otolith input plays a key role in supporting the brain’s internal representation of verticality and spatial proportions.

## Discussion

These findings indicate that removing visual input significantly alters spatial output in vestibular patients, leading to vertical compression of drawn and written figures. This supports the hypothesis that vestibular input is critical for maintaining accurate spatial metrics in the absence of vision. The vertical compression of spatial output while in microgravity mirrors the distortions seen in vestibular patients tested without vision. The consistency of this effect across both populations—vestibular-impaired patients on Earth and healthy individuals in the altered sensory environment of space—strongly suggests that otolith input plays a key role in supporting the brain’s internal representation of verticality and spatial proportions.

In vestibular patients, the vertical-to-horizontal length ratio was, on average, 10.7 ± 2.2% lower with eyes closed compared to eyes open, across all shapes. This finding aligns with a previous study in which patients were shown line drawings of squares and circles with vertical dimensions that were equal to, 5-10% shorter, or 5-10% taller than their width. For each shape, participants were asked to judge whether it appeared “perfect” or “not perfect.” Otolith patients more frequently judged shapes with a 5-10% taller height as “perfect”, and often perceived true 1×1 shapes as appearing too short [7].

In astronauts, the vertical-to-horizontal length ratio was, on average, 9.9 ± 2.5% lower during spaceflight compared to preflight measurements, across all shapes. This is consistent with findings from a separate in-flight study, where astronauts viewed a stereoscopic image of a cube presented in perspective. One of the cube’s dimensions (width, height, or depth) was intentionally distorted—either clearly shorter or longer than the others—and participants were asked to adjust it to match the apparent size of the remaining two. After 3 months in microgravity, astronauts consistently adjusted the height to be approximately 10% shorter than the width, indicating a systematic underestimation of the vertical dimension in space [6].

In another study, two astronauts drew ellipses in microgravity, aligned either parallel (longitudinal) or perpendicular (transverse) to the body axis. Consistent with our findings, the vertical lengths of longitudinal ellipses decreased by 11.7% and 12.8% during flight, and transverse ellipses showed vertical reductions of 4.0% and 10.1%. In contrast, horizontal lengths changed by only ±2 ± 4% inflight compared to before the flight [8,9].

Vestibular patients with otolithic dysfunction often report distortions in the perceived size of their body and surrounding objects, especially along the vertical axis. For example, people, cars, or buildings may appear abnormally small, or the ground may seem too close—a condition known as micropsia [10]. Neuropsychological evidence also supports the otolith system’s role in body schema. For example, vestibular stimulation can temporarily correct spatial neglect and body image distortions in brain-damaged patients [11]. Similarly, individuals with somatosensory deafferentation (loss of proprioceptive and tactile input) show distortions in drawing tasks, reflecting a disrupted body schema [12]. Like proprioceptive loss, vestibular dysfunction disrupts the sensorimotor framework the brain uses to orient the body in space [13]. Both systems deliver continuous, unconscious input essential for maintaining a coherent perception of the self within the environment. The idea that spatial perception is distorted in astronauts and vestibular patients with otolith dysfunction is further supported by their reduced susceptibility to geometric illusions based on perspective [7,14,15].

The otolith organs are essential for sensing orientation relative to gravity, which plays a central role in how we perceive the world. On Earth, visual cues like the horizon or falling objects help define “up” and “down,” supporting internal models that guide spatial perception and cognition [16]. In microgravity, these cues disappear—objects and bodies float freely, and the sense of verticality becomes irrelevant. Astronauts lose proprioceptive input when free-floating, while vestibular patients rely heavily on proprioception to compensate for vestibular loss. Both vestibular and proprioceptive signals are crucial for calibrating motor output, ensuring appropriate movement size and force when drawing and writing. When vestibular and visual inputs are unreliable, the brain struggles to maintain accurate spatial metrics, contributing to size distortions in drawings and altered spatial cognition, including perceptions of body and object size.

Taken together, these results indicate that both visual and otolith cues contribute to the accurate scaling of motor output in drawing and writing tasks. When either input is absent or degraded, the brain’s internal spatial representation shifts—leading to systematic distortions, particularly in the vertical dimension. This parallel between clinical and spaceflight conditions underscores the importance of vestibular function in spatial cognition and motor planning.

The connection between vestibular and cognitive functions likely stems from direct neural pathways linking vestibular signals to cortical areas involved in spatial processing. The parietoinsular vestibular cortex, along with the posterior parietal cortex, plays a key role in integrating vestibular input for spatial awareness [17]. Alterations in drawing size may serve as subtle indicator of otolith dysfunction. When evaluated alongside orientation and shape distortions, they can enhance the sensitivity of clinical assessments. These size changes may also reflect broader functional impairments, such as difficulties with navigation and postural control.

## Data Availability

All data produced in the present work are contained in the manuscript.

